# Forecasting sub-national trends in COVID-19 vaccine uptake in the UK

**DOI:** 10.1101/2020.12.17.20248382

**Authors:** A. de Figueiredo

## Abstract

The rollout of COVID-19 vaccines has begun to at-risk populations around the world. It is currently unclear whether rejection of the vaccine will pose challenges for achieving herd/community immunity either through large-scale rejection or localised pockets. Here we predict uptake of the vaccine at unprecedented spatial resolution across the UK using a large-scale survey of over 17,000 individuals. Although the majority of the UK population would likely take the vaccine, there is substantial heterogeneity in uptake intent across the UK. Large urban areas, including London and North West England, females, Black or Black British ethnicities, and Polish-speakers are among the least accepting. This study helps identify areas and socio-demographic groups where vaccination levels may not reach those levels required for herd immunity. Identifying clusters of non-vaccinators is extremely important in the context of achieving herd immunity as vaccination “cold-spots” can amplify epidemic spread and disproportionately increase vaccination levels required for herd protection.

## Introduction

A vaccine against the novel coronavirus 2019 disease (COVID-19) caused by the severe acute respiratory coronavirus 2 (SARS-CoV-2) will be a major step in reducing mortality, morbidity, economic, and societal burdens associated with the COVID-19 pandemic. The UK’s National Health Service (NHS) has begun the rollout of two vaccines approved by the Medicines and Healthcare products Regulatory Authority (MHRA)^1^ and has administered almost 4 million doses (week ending 17 January 2021).

A successful vaccination campaign of a safe and effective COVID-19 vaccine is contingent on several factors: at-scale manufacture ensuring sufficient dosages to target populations; governments and health organisations ensuring fast and equitable distribution via existing and novel supply-chain networks with sufficient capacity for storage and delivery; and public acceptance. This latter factor is perhaps of particular concern in the UK, which has had notable hesitancy towards vaccinating in the past^2^, and has had widely circulating false stories about a COVID-19 vaccine^3–5^. Over the past three years, there have been year-on-year decreases in uptake of routine immunisations – such as the MMR vaccine^6^ – with corresponding outbreaks of vaccine-preventable diseases (and a loss of the UK’s measles-free status).^7–10^. UK policymakers may therefore face significant public concern over a novel vaccine.

Although recent studies show largely positive attitudes towards a COVID-19 vaccine across the UK^11,12^, no study has – to date – investigated sub-national acceptance and whether specific regions may fail to meet the required vaccination levels for herd immunity (estimated at around 65% for the UK^13^). Vaccine delays and refusals not only place individuals directly at risk but can contribute to lowering vaccination thresholds required for herd immunity. Geographic clustering of non-vaccinators can be particularly troublesome, as these “cold spots” can disproportionately increase required vaccination levels for herd immunity in adjacent settings, as they serve as infection hubs amplifying the spread of disease^14,15^. It is therefore important to identify the regions – and the socio-demographic groups – at risk of vaccine refusal or delay.

In this large-scale modelling study, intent to accept a COVID-19 vaccine is estimated for 174 sub-national regions across the UK using survey data from over 16,820 individuals. Multilevel regression and poststratification (MRP) – a statistical method recently used to successfully predict national general election results in the UK^16^ – is used to obtain these sub-national estimates and to identify the socio-demographic barriers of intent to accept a COVID-19 vaccine. Partial validation for this modelling approach is obtained via uptake rates among over 70s across England from the start of vaccine rollout to 18 February 2021.

This study aims to provide policymakers with estimates for COVID-19 uptake rates among the UK adult population and to establish socio-demographic groups at high risk of vaccine refusal and to highlight regions that may pose challenges for reaching herd immunity across the UK.

## Results

### Data collection

Between 24 September and 14 October 2020, a cross-sectional online survey was administered to 17,684 UK residents aged 18 and over. During data collection, quality control procedures resulted in the removal of 864 respondents (see *Methods*). All respondents were recruited via an online panel by ORB (Gallup) International (www.orb-international.com) and informed consent was obtained before respondents participated. The questionnaire is provided in the Supplementary Materials.

Respondents are asked whether they would accept a COVID-19 vaccine: “*If a new coronavirus (COVID-19) vaccine became available, would you accept the vaccine for yourself?*”. Respondents could provide one of four responses on an ordinal scale: “*yes, definitely*”, “*unsure, but leaning towards yes*”, “*unsure, but leaning towards no*”, or “*no, definitely not*”.

Socio-demographic data was collected for each respondent to assess the relationship between these characteristics and vaccine intent and to allow for the reweighting of respondents’ vaccination intent according to census data (both via multilevel regression and poststratification, see below). These covariate data were therefore chosen to align with the socio-demographic data collected in the latest UK census. The covariate data collected for each individual was: sex, age, highest educational attainment, religious affiliation, ethnicity, employment status, primary language, and outer postcode. Respondent’s outer postcode was used to map respondents to one of 174 third level NUTS regions (NUTS3). Descriptions for all respondent data collected and recoding are provided in table 1.

**Table 1.**
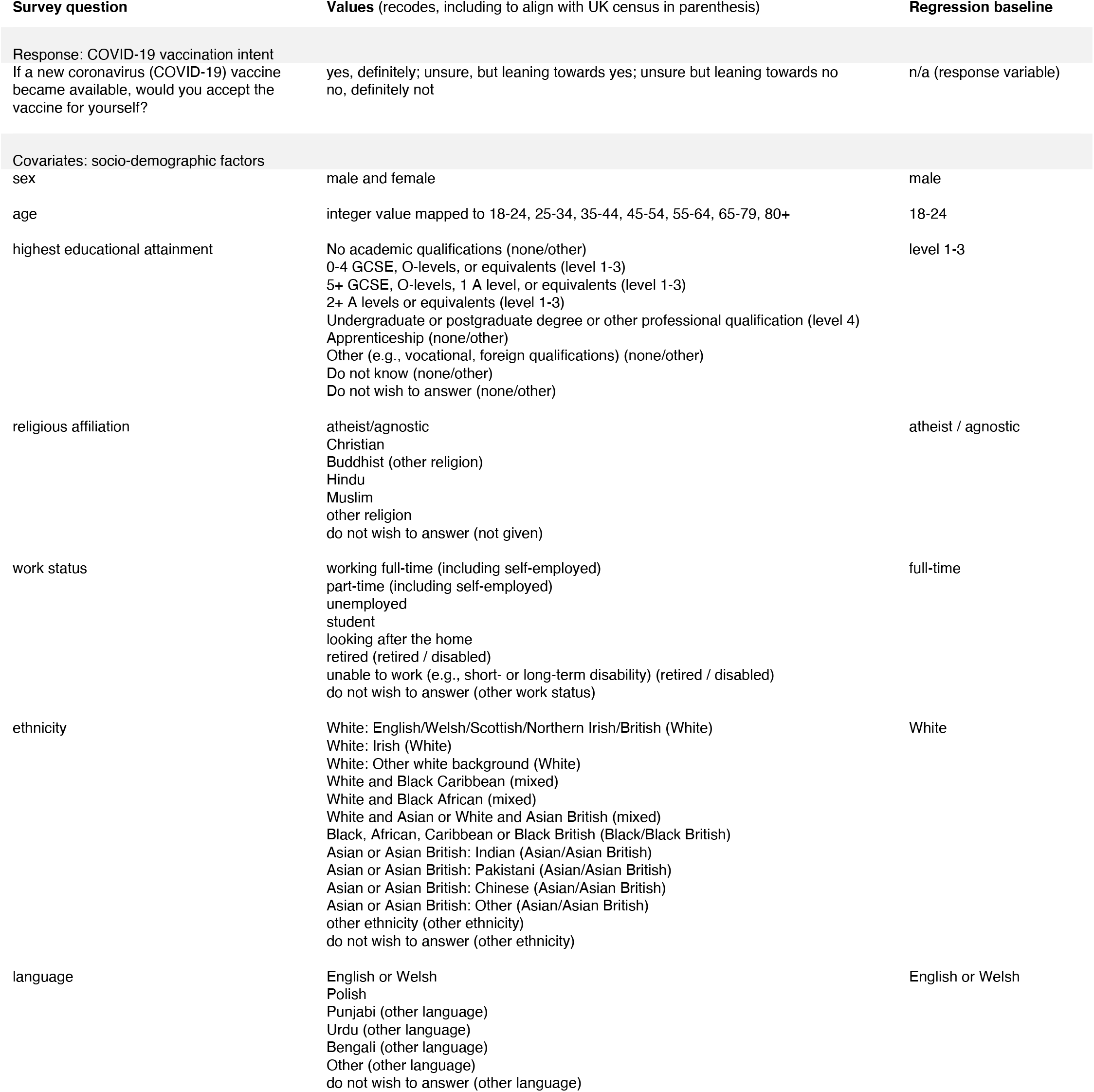
Study data. Survey items are shown with the responses (including recodes, if any), and baselines used in the ordinal logistic regressions. COVID-19 vaccination intent is the study response variable.

| Survey question | Values (recodes, including to align with UK census in parenthesis) | Regression baseline |
| --- | --- | --- |
| Response: COVID-19 vaccination intent<br>If a new coronavirus (COVID-19) vaccine became available, would you accept the vaccine for yourself? | yes, definitely; unsure, but leaning towards yes; unsure but leaning towards no<br>no, definitely not | n/a (response variable) |
| Covariates: socio-demographic factors |  |  |
| sex | male and female | male |
| age | integer value mapped to 18-24, 25-34, 35-44, 45-54, 55-64, 65-79, 80+ | 18-24 |
| highest educational attainment | No academic qualifications (none/other)<br>0-4 GCSE, O-levels, or equivalents (level 1-3)<br>5+ GCSE, O-levels, 1 A level, or equivalents (level 1-3)<br>2+ A levels or equivalents (level 1-3)<br>Undergraduate or postgraduate degree or other professional qualification (level 4)<br>Apprenticeship (none/other)<br>Other (e.g., vocational, foreign qualifications) (none/other)<br>Do not know (none/other)<br>Do not wish to answer (none/other) | level 1-3 |
| religious affiliation | atheist/agnostic<br>Christian<br>Buddhist (other religion)<br>Hindu<br>Muslim<br>other religion<br>do not wish to answer (not given) | atheist / agnostic |
| work status | working full-time (including self-employed)<br>part-time (including self-employed)<br>unemployed<br>student<br>looking after the home<br>retired (retired / disabled)<br>unable to work (e.g., short- or long-term disability) (retired / disabled)<br>do not wish to answer (other work status) | full-time |
| ethnicity | White: English/Welsh/Scottish/Northern Irish/British (White)<br>White: Irish (White)<br>White: Other white background (White)<br>White and Black Caribbean (mixed)<br>White and Black African (mixed)<br>White and Asian or White and Asian British (mixed)<br>Black, African, Caribbean or Black British (Black/Black British)<br>Asian or Asian British: Indian (Asian/Asian British)<br>Asian or Asian British: Pakistani (Asian/Asian British)<br>Asian or Asian British: Chinese (Asian/Asian British)<br>Asian or Asian British: Other (Asian/Asian British)<br>other ethnicity (other ethnicity)<br>do not wish to answer (other ethnicity) | White |
| language | English or Welsh<br>Polish<br>Punjabi (other language)<br>Urdu (other language)<br>Bengali (other language)<br>Other (other language)<br>do not wish to answer (other language) | English or Welsh |

### Multilevel regression and poststratification

Multilevel regression and poststratification (MRP)^16,17^ is used to estimate intent to accept a COVID-19 vaccine across the 174 sub-national regions across the UK and to identify the socio-demographic barriers to uptake (see *Methods* for full model details).

### COVID-19 vaccination intent

Across the UK, just under half the population – 47.5% (95% highest posterior density interval (HPDI) 46.5 to 48.5%) – would “definitely” take a COVID-19 vaccine according to the MRP-based estimates of uptake intent. A further 32.6% (31.8 to 33.2%) are leaning towards vaccinating but are unsure. 8.7% (8.2 to 9.2%) would “definitely not” take a COVID-19 vaccine and 11.2% (10.7 to 11.8%) are unsure but leaning towards no (fig 1).

**Fig. 1.**
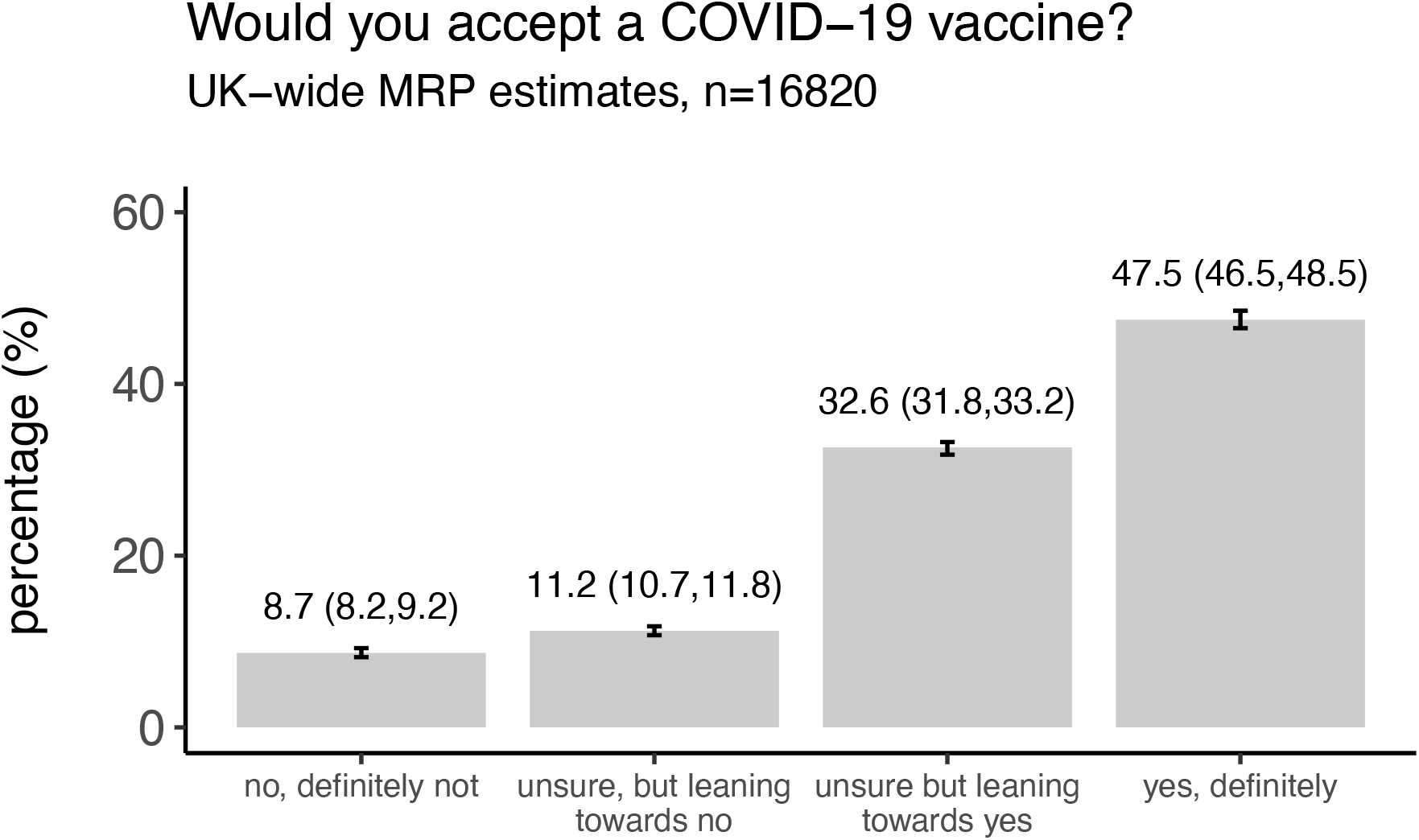
National-level estimates of COVID-19 vaccine uptake intent. National-level estimates for the percentage of the UK that would: “definitely not” accept a COVID-19 vaccine, “definitely” accept a COVID-19 vaccine, or who are unsure. Uncertainty in estimates are 95% HPD intervals.

Sub-national MRP estimates of the proportion of each of the UK’s 174 NUTS regions who would “definitely” accept a COVID-19 vaccine are mapped in figure 2A. Estimates of the proportions who would “definitely not” accept a COVID-19 vaccine are mapped in figure 2B. The values in figure 2A are repeated in figure 3 with their corresponding 70% and 95% HPDIs and are ranked from lowest to highest acceptance by broad UK region. (Raw values for the MRP estimates and HPDIs for each of the 174 sub-national regions and each of the four outcome variable options are provided in the supplementary data file).

**Fig. 2.**
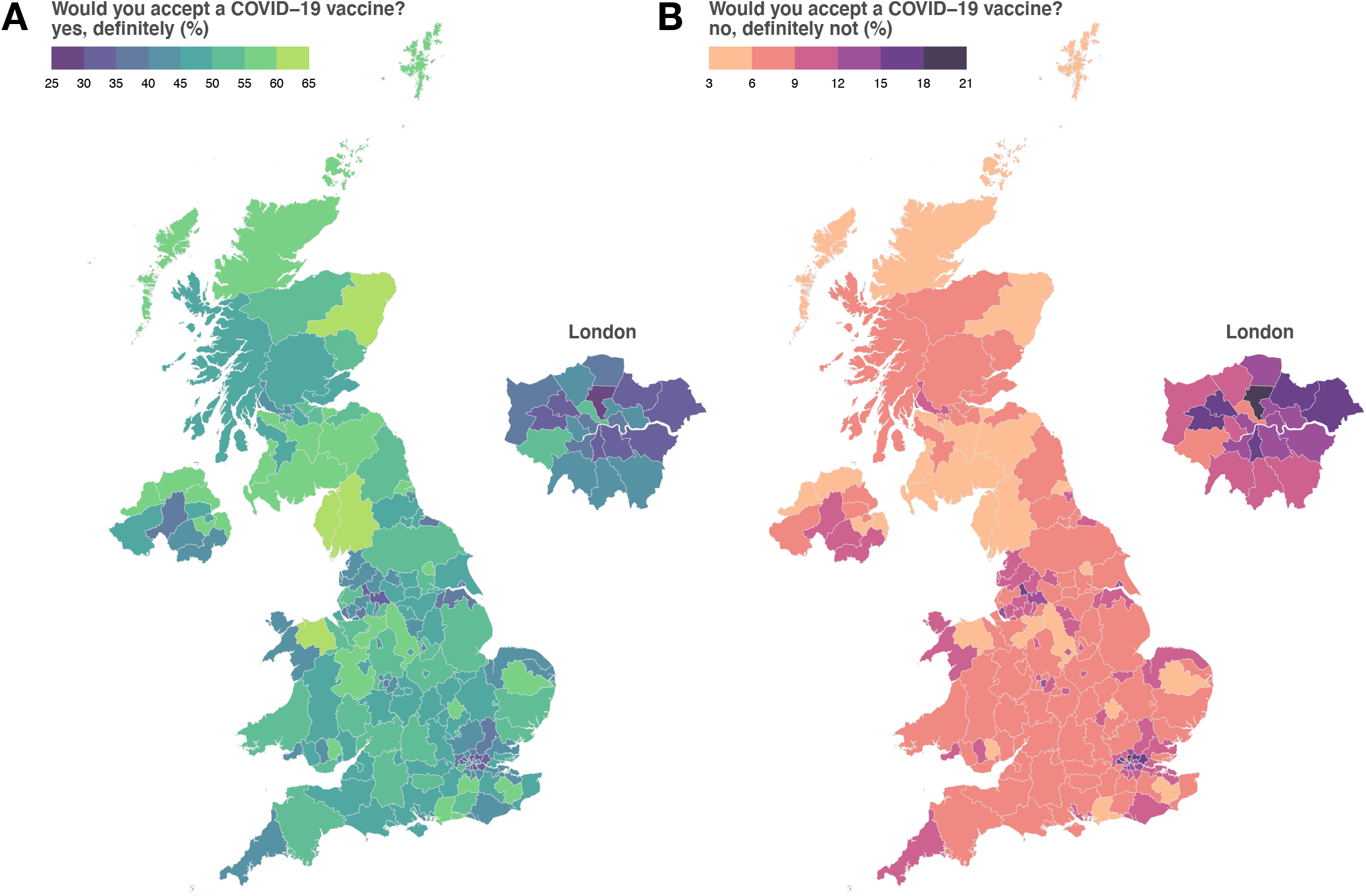
Sub-national estimates of COVID-19 vaccine intent across the UK. The estimated proportion of respondents in each of the UK’s 174 NUTS3 region who would state they would “definitely” accept a COVID-19 (A) and who would “definitely not” accept a COVID-19 vaccine (B). Regional boundaries are used under the Open Government License v3.0 (see https://data.gov.uk/dataset/b147a160-86b6-48e4-8dd0-f35b90981814/nuts-level-3-january-2015-super-generalised-clipped-boundaries-in-england-and-wales accessed 25 November 2020).

**Fig. 3.**
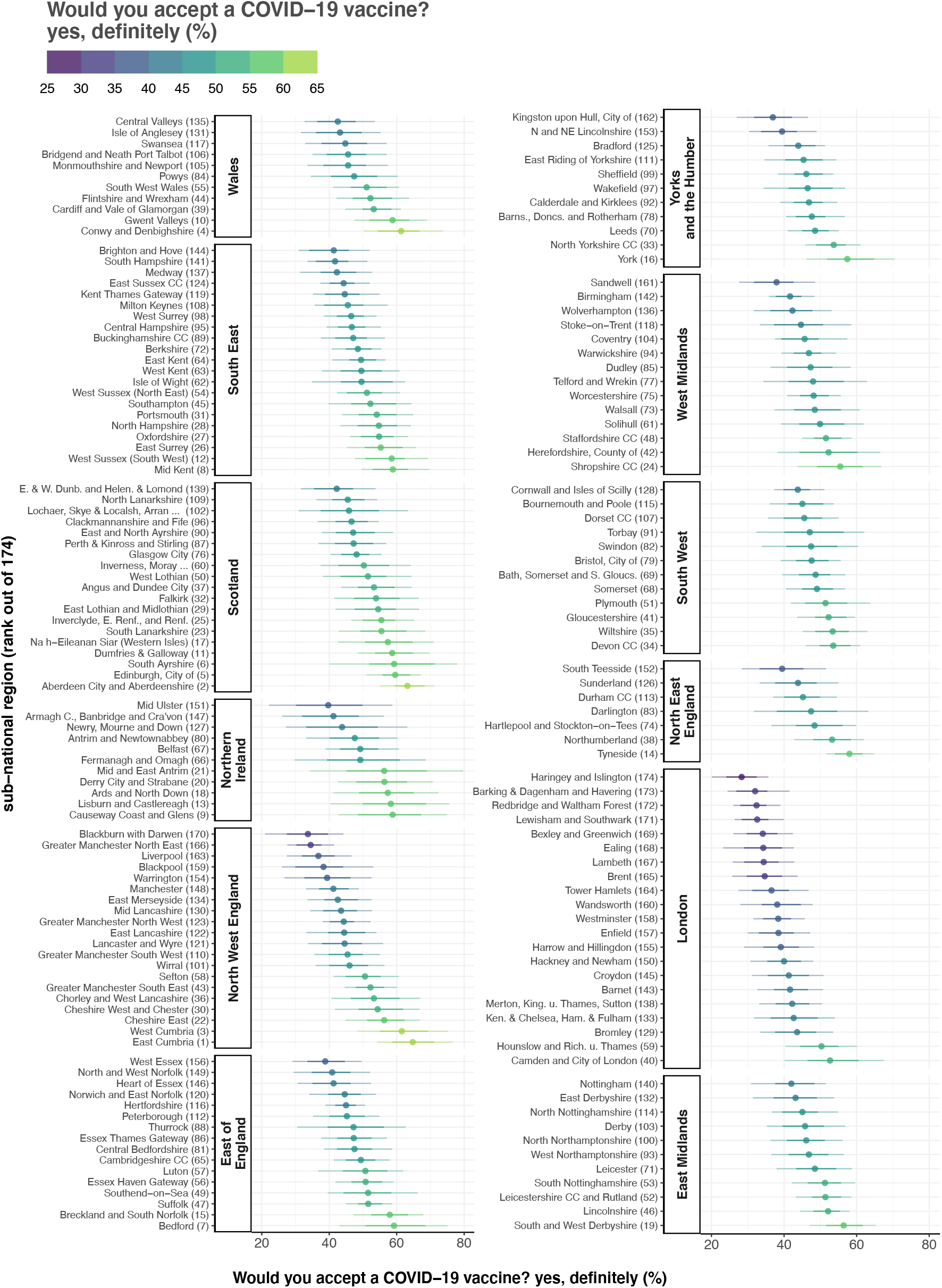
Ranked intent to accept a COVID-19 vaccine. The estimated proportion of respondents in each of the UK’s 174 NUTS3 region who would definitely accept a COVID-19 vaccine are shown and ranked within the 12 first-level NUTS regions (NUTS1). 70% and 95% highest posterior density intervals (horizontal bars) are shown around the mean estimate (dot). Each region is suffixed by its rank (out of 174) according to the estimated proportion who would “definitely” accept a COVID-19 vaccine. East Cumbria (North West England) ranks first, while Haringey and Islington (London) is last.

Estimates across the 174 sub-national NUTS regions of the UK vary considerably. Estimates of the proportion of the public who would “definitely” accept a COVID-19 vaccine (figure 2A) range from 28.3% (20.1 to 35.7%) in Haringey and Islington to 64.8% (54.2 to 76.6%) in East Cumbria (fig 2 and fig 3). The lowest proportions of the UK public who would “definitely” accept a COVID-19 vaccine are concentrated in London, which contains 13 of the 20 lowest proportions in the UK: Haringey and Islington (28.3%, 20.1 to 35.7%); Barking, and Dagenham and Havering (32.0%, 24.4 to 41.5%); Redbridge and Waltham Forest (32.4%, 26.0 to 39.1%); Lewisham and Southwark (32.6%, 26.4 to 39.9%); Bexley and Greenwich (34.1%, 26.1 to 42.4%); Ealing (34.3%, 23.2% to 42.7%); Lambeth (34.4%, 25.8 to 42.8%); Brent (34.7%, 25.7 to 43.7%); Tower Hamlets (36.5%, 27.4 to 46.8%); Wandsworth (38.1%, 28.0 to 47.9%); Westminster (38.4%, 31.6 to 45.7%); Enfield (38.5%, 30.0 to 47.2%); and Harrow and Hillingdon (39.1%, 29.0 to 48.3%). Four of the remaining seven regions in the lowest 20 are in North West England: Blackburn with Darwen (33.7%, 21.0 to 44.2%), Greater Manchester North East (Bury, Oldham, and Rochdale) (34.5%, 27.5 to 41.4%), Liverpool (36.8%, 27.4 to 46.6%), Blackpool (38.3%, 25.9 to 53.0%). The remaining three areas in the lowest 20 are West Essex (East of England, 38.8%, 29.1 to 49.6%), Sandwell (West Midlands, 37.9%, 27.6 to 48.6%), and the City of Kingston upon Hull (Yorkshire and the Humber, 36.9%, 27.0 to 46.6%). (See supplementary data file for all estimated values and posterior intervals.)

The five regions with the highest proportions of the UK public who would “definitely” accept a COVID-19 vaccine are East Cumbria (64.8%, 54.2 to 76.6%), Aberdeen City and Aberdeenshire (63.2%, 54.9 to 71.1%), West Cumbria (61.5%, 48.3 to 75.3%), Conwy and Denbighshire (61.3%, 50.3 to 73.7%), and the City of Edinburgh (59.6%, 51.0 to 67.1%). The top 20 regions disproportionately contain regions in Scotland (5 regions) and Northern Ireland (4) (fig 3 and the supplementary data file). (In fact, Scotland and NI have 13 of the highest-ranking regions in the top 26.)

The regions with the highest estimated proportions who would “definitely not” accept a COVID-19 vaccine are again predominately located in London and the North West. Haringey and Islington (19.0% (12.8 to 25.6%), Blackburn with Darwen (16.6, 9.8 to 25.0), and Redbridge and Waltham Forest (16.3%, 11.7 to 21.1%) have the highest estimated proportions who would “definitely not” accept the vaccine, while East Cumbria (3.8%, 2.0 to 5.8%), Aberdeen City and Aberdeenshire (4.1%, 2.8 to 5.9%), and West Cumbria (4.5%, 2.2 to *7*.5%) have the lowest. Estimates for the proportions of respondents who are “unsure” about taking a COVID-19 vaccine are provided in the supplementary data file and mapped in the appendix, figure S1.

### Socio-demographic determinants of vaccination intent

The fixed-effects in the ordinal multilevel regression (see statistical analysis and appendix 2) – which represent an “average” impact of socio-econo-demographic characteristics on vaccination intent across the whole country – are shown in figure 4.

**Fig. 4.**
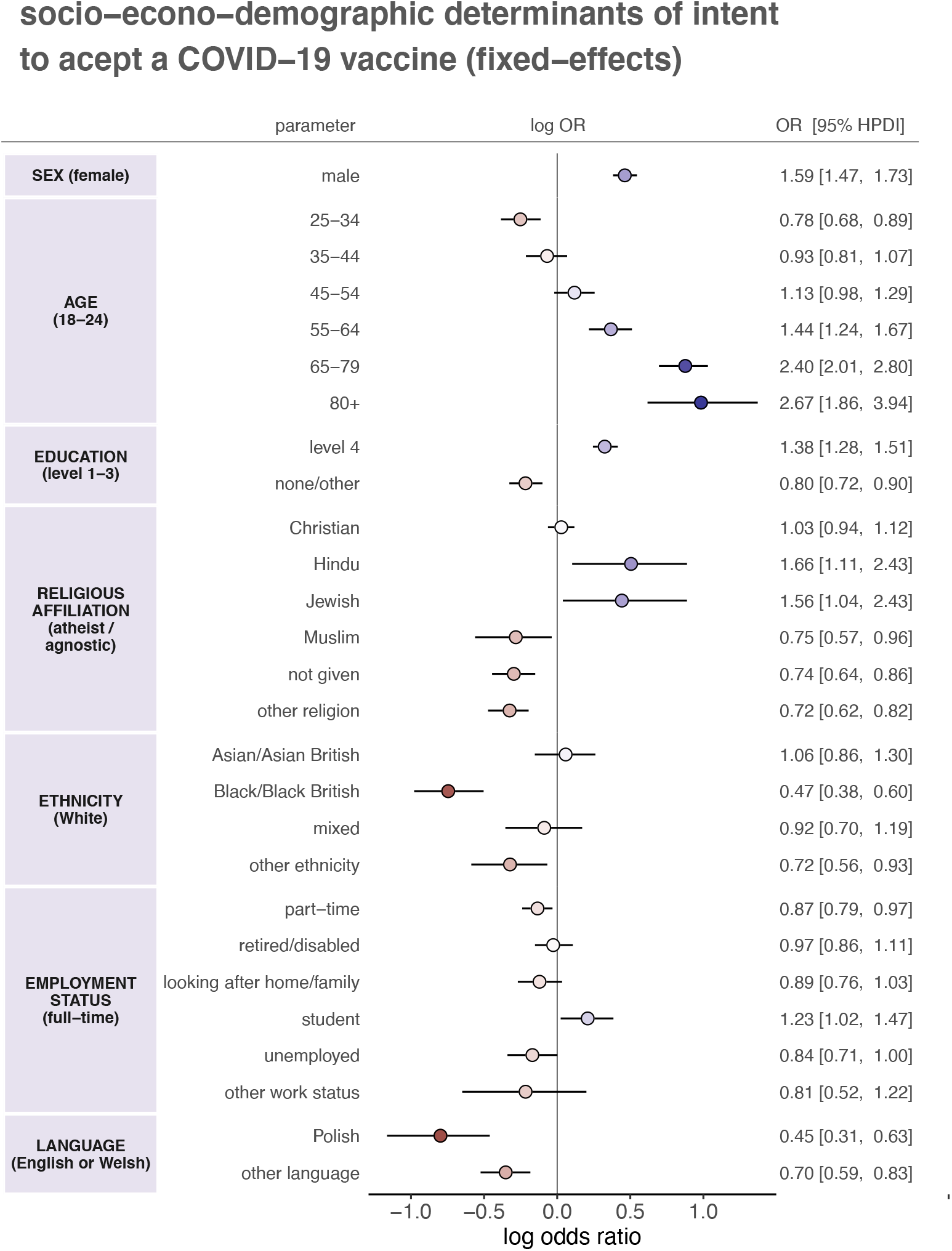
The socio-demographic determinants of intent to accept a COVID-19 vaccine. Multilevel regression fixed-effect parameter log odds ratios are plotted with corresponding 95% HPDIs. These log odds ratios are coloured by effect magnitude and direction, where blues (reds) signify that the group is more (less) likely than the baseline group to accept a COVID-19 vaccine. The darker the colour the stronger the association. For each factor, the baseline group is provided in parentheses on the left. Odds ratios with 95% HPDIs are shown on the right for each parameter.

A number of factors are associated with COVID-19 vaccine intent. Males are more likely than females (odds ratio 1.59, 95% HPDI 1.47 to 1.73) to accept a COVID-19 vaccine. Older age groups are more likely to accept a COVID-19 vaccine than 18-24-year-olds, in particular 65-79 and 80+ year-olds (2.40, 2.01 to 2.80 and 2.67, 1.86 to 3.94, respectively). Interestingly, 25-34-year-olds are *less* likely than 18-24-year-olds to accept a vaccine (0.78, 0.68 to 0.89). Individuals with undergraduate or postgraduate qualifications (level 4) are more likely than those with GCSEs, A- or O-levels to accept a vaccine (1.38, 1.28 to 1.51) while those with no formal qualifications or other qualifications (see table 1) are less likely (0.80, 0.72 to 0.90).

There is no evidence to suggest those who identify as Christian are more or less likely than atheists or agnostics to accept a vaccine (1.03, 0.94 to 1.12), those reporting Hinduism or Judaism as their religion are more likely than atheists or agnostics to accept a vaccine (1.66, 1.11 to 2.43 and 1.56, 1.04 to 2.43, respectively). Those identifying as Muslim (0.75, 0.57 to 0.96), not providing their religion (0.74, 0.64 to 0.86), or stating an “other” religious affiliation (0.72, 0.62 to 0.82) are less likely to accept a COVID-19 than atheists or agnostics. Ethnicity also plays a role in determining intent to accept a COVID-19 vaccine, independently of religion, with those identifying as Black or Black British (0.47, 0.38 to 0.60) and those reporting an “other” ethnicity than those provided (0.72, 0.56 to 0.93) less likely to accept a COVID-19 vaccine than Whites.

Individuals’ employment status appears to play less of a role than the other factors outlined above, with odds ratios closer to one. However, there is evidence to suggest that those in part-time work (0.87, 0.79 to 0.97) or who are unemployed (0.84, 0.71 to 1.00) are less likely than those in full-time in employment to accept a COVID-19 vaccine, while students (1.23, 1.02 to 1.47) are more likely.

Individuals who report a language other than English or Welsh as their primary language hold less intent to accept to accept a vaccine than those reporting English or Welsh (Polish 0.45, 0.31 to 0.63 and “other” language 0.70, 0.59 to 0.83).

Variation in socio-demographic determinants of uptake across the UK are shown in figure 4 for the fixed-effect parameters with the strongest overall association with uptake intent. To focus on the strongest associations between socio-demographic factor and uptake, regions are coloured if the 95% HPDI excludes zero and set to zero otherwise. Males are found to be more likely to accept a COVID-19 vaccine in 67 UK regions (figure 5A), while 65–79-year-olds are found to be more likely to accept the vaccine than 18–24-year-olds in all but eight UK regions (figure 5B). There is evidence that individuals identifying as Black/Black British are less likely than those identifying as White to accept the vaccine in 17 UK regions mostly concentrated in London (e.g., Hackney and Newham, Tower Hamlets, Haringey and Islington, Lewisham and Southwark), the West Midlands (Birmingham Sandwell, and Wolverhampton), and Brighton and Hove (figure 5C). Polish speakers are found to be less likely than English or Welsh speakers to accept the vaccine in seven UK regions (figure 5D). All region-specific random-effect parameters with corresponding HPD intervals are provided for national and local policymakers in the supplementary data file and shown in SM figure S3.

**Fig. 5.**
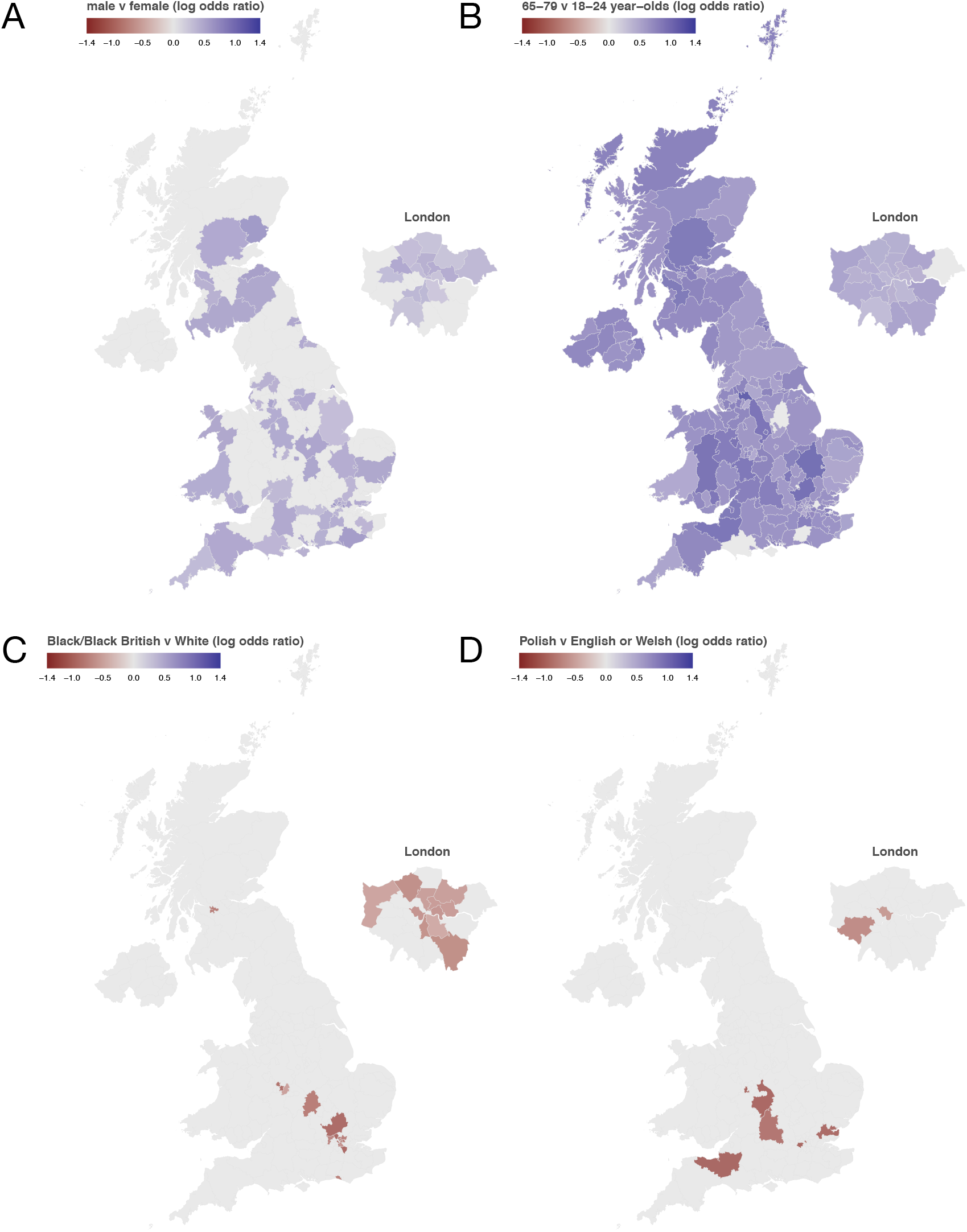
Sub-national socio-demographic determinants of intent to accept a COVID-19 vaccine. Multilevel regression random-effect parameter log odds ratios are shown for the four socio-demographic sub-groups that show the strongest differential association with uptake. Log odds ratios are shown for each sub-national region for A) sex (males versus females), B) age (65-79 year-olds versus 18-24 year-olds), C) ethnicity (Black/Black British v White), D) language (Polish v English or Welsh). Blues denote that the group has a positive association with intent to accept a COVID-19 vaccine with respect to the baseline group, while holding all other covariates constant. For example, in all cases when the 95% HPDI intervals around the male random-effect parameters exclude zero, males are more likely than females to state they would accept the vaccine. Parameters whose 95% HPDIs exclude zero (all other parameters are set to zero for the purpose of visualisation)

### Model validation against recent UK uptake data

Vaccination rollout began in the UK on 8 December 2020. Data are now available on the percentage of first doses administered in each of England’s 136 Clinical Commissioning groups (CCG) for over 70s (who have now been offered a vaccine) as well as the estimated over 70 population in each of these regions^18^. To obtain validation for the modelling approach in this study, two checks are performed. Firstly, all regional forecasts across England are correlated with observed uptake among over 70s and secondly, multilevel regression and poststratification is reimplemented for all individuals aged over 65 (census microdata records bin respondents by age, and so it was necessary to include some respondents less than 70 so as to not remove respondents aged 70-74 from the analysis) at the second NUTS level. As there are 3,338 individuals aged over 65 collected in the survey, MRP estimates are generated at the 40 second level NUTS units across the UK (of which 33 units are in England). Occasionally, regional boundaries do not precisely align between CCG and NUTS units and, when this is the case, population-weighted averages are taken to provide a reasonable mapping between acceptance forecasts in the NUTS units and the CCGs. First dose data are used, since the UK vaccination policy involves a four-to 12-week delay between of either the Pfizer-BioNTech or Oxford-AstraZeneca vaccine in the majority of cases, and will therefore be more representative of uptake compared to a completed two-dose schedule^19^.

Predicted vaccine acceptance (the percentage of respondents stating they would “definitely” vaccinate or who are “unsure, but leaning towards yes”) across all England regions and for all adults correlates with observed uptake among over 70s (*ρ*=0.53 (0.37, 0.62), figure 6A). Predicted uptake is lower than observed uptake, because COVID-19 vaccines have not been made available to the vast majority of younger age groups. Predicted acceptance among all over 65s surveyed also correlates with observed uptake among over 70s, though this correlation is weaker (*ρ*=0.31 (0.00, 0.58), figure 6B). Predicted uptake is again lower than observed, which is possibly due to 65–69-year-olds being included in the analysis or because of changes in the UK’s vaccine acceptance since survey data was collected in late 2020.

**Fig. 6.**
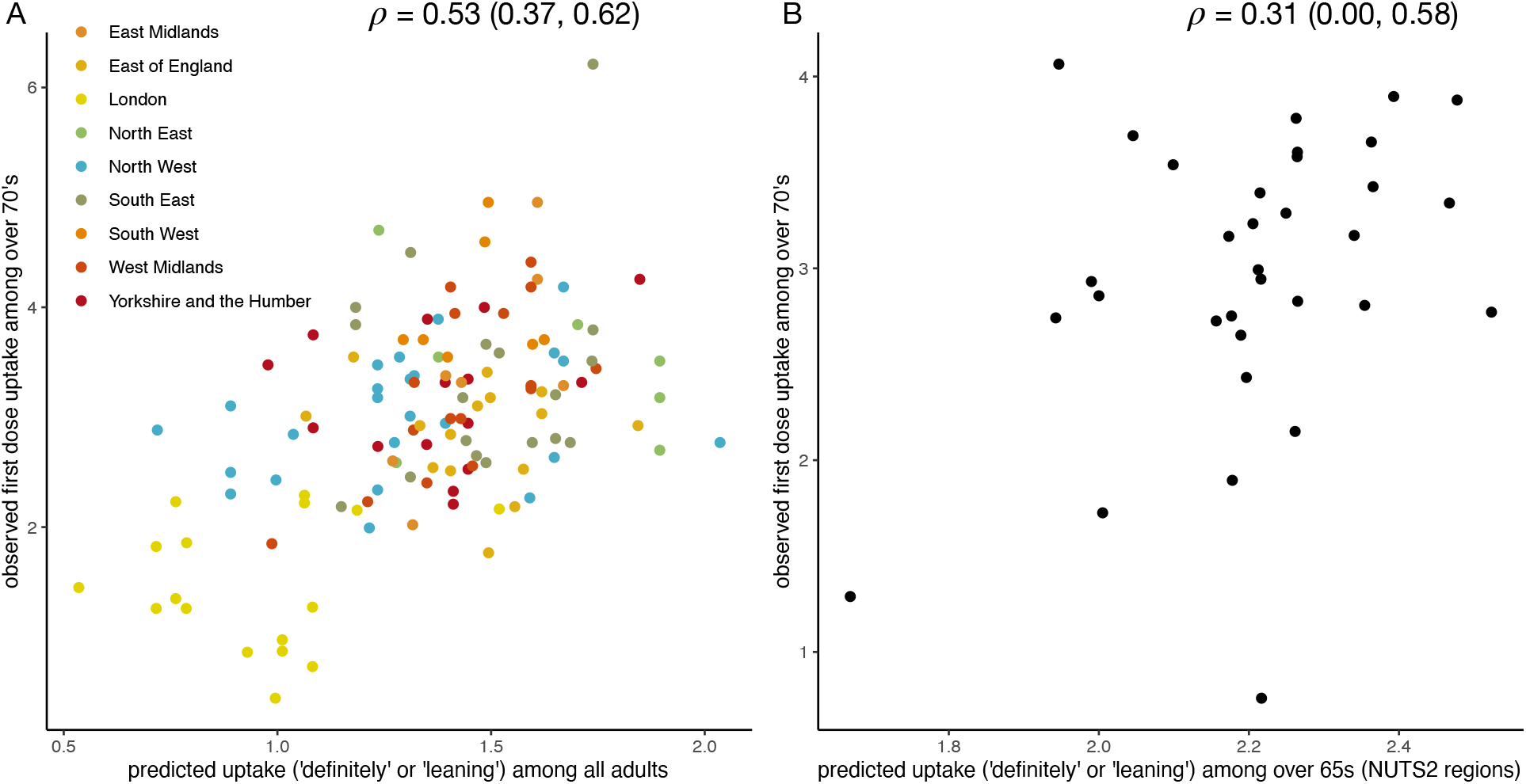
Forecasts of vaccine acceptance correlates at sub-national levels correlates with observed first-dose COVID-19 vaccine uptake among over 70s. (A) Observed vaccination coverage across all England Clinical Commissioning Groups correlates with predicted uptake across the UK adult popualtion. (B) Obseverd vaccination coverage across 33 NUTS2 levels across England correlates with predicted uptake among over 65s surveyed in this study. Coverage values on both axes have been scaled using the inverse logit transform.

## Discussion

This study reports multiple findings of immediate relevance to clinicians and policymakers involved with the delivery of a COVID-19 vaccine. This study estimates that less than half the UK public would “definitely” accept a COVID-19 vaccine, with strong regional variation in estimates. Although a relatively small proportion (8.7%, 8.2 to 9.2%) state that they would “definitely, not” accept a vaccine, rates of rejection intent are much higher in London and the North West, where they reach as high as 18.0% (14.8 to 20.7%) in Haringey and Islington. Since February 2020, London and the North West have experienced high disease burdens. The North West is particularly notable in this regard as four of the five UK regions with the highest infection rate (correct of 20 November 2020) are all in the North West: Blackburn and Darwen (6,312 per 100,000), Oldham (6,157), Rochdale (5,585), and Manchester (5,539)^20^. Interestingly, Blackburn and Darwen has the fifth lowest intent to accept a COVID-19 vaccine (ranking 170 out of 174), while Oldham and Rochdale rank ninth lowest (Greater Manchester North East – which contains both these towns – ranks 166 out of 174). Manchester fares a little better ranking 148 out of 174. These results point to an important possible interaction between high COVID-19 rates and low vaccine acceptance and the effect this may have on vaccination rates required for herd/community immunity in adjacent regions^13–15^. Significant correlations are observed between sub-national forecasts and first-dose vaccination uptake among over 70s, validating the modelling approach.

Socio-demographic background is strongly associated with intent to accept the vaccine. This study finds strong evidence to suggest that males, and older age groups are substantially more likely to accept a COVID-19 vaccine than females and 18-24-year-olds (respectively). Highest level of education, ethnicity, religious affiliation, and primary language are also found to be strongly related to intent to accept a COVID-19 vaccine. Most notably, individuals who identify as Black or Black British are much less likely than Whites to intend to receive the vaccine, as too are those reporting Polish as their primary language. These associations have been found with regards to existing immunisation programmes^21–24^, as well as – more recently – with respect to vaccine acceptance of a COVID-19 vaccine specifically. A study of over 30,000 adults in the UK conducted between 7 September to 5 October, found similar rates of intent to reject a vaccine (14% of respondents unwilling to receive a vaccine compared to 8.7% -- comparison of intent to accept a vaccine is difficult due to differences in questionnaire wording and socio-demographic drivers of intent^12^. Notably, that females and those with education levels below postgraduate degrees were less likely to accept a COVID-19 vaccine. A link between BAME groups and uptake was not found at a 95% significance testing interval, however^12^ (this could be because of the aggregation of BAME groups and/or a different set of predictor variables used to explain variation in uptake intent). Two other recent studies examining COVID-19 vaccine intent in England and Scotland, however, do find that intent to accept a COVID-19 vaccine is modulated by ethnicity, with non-Whites less likely to accept a COVID-19 vaccine^25,26^. As risk of severe COVID-19 is greater in BAME communities^27^, achieving high vaccine acceptance may avert further burden within these communities. (The author refers policymakers to the supplementary data file which reveals regions in which there is a strong association between ethnicity and uptake intent.)

There are a number of study limitations to note. Firstly, this study maps intent to accept a COVID-19 vaccine across the entire population and does not assess vaccine acceptance among at-risk groups or healthcare workers, who are likely to be the first groups offered a novel vaccine. Secondly, the most recent census data used for probability reweighting (see *Statistical analysis* and appendix 2) is from 2011. Large changes in the demographic structure of the 174 regional populations could, therefore, result in biased estimates of vaccine intent. Finally, the study was conducted online with a sample of panellists who registered to take part in research surveys. While efforts have been made to ensure representativeness via MRP, there may be a bias among respondents who have access to (and can use) mobile phones or computers, through which the questionnaire would be completed.

While this study provides a comprehensive snapshot of intent to accept a vaccine across the UK in September and October 2020, it predates both the Pfizer announcement that approval is being sought for use in the UK and the peak of the second wave of daily new coronavirus cases. Attitudes may change on short timescales. As the second wave passes, the UK public may have a decreased appreciation for the importance of the vaccine through either a decrease in the perception of the seriousness of disease or a belief that they have already been infected with SARS-CoV-2 (which is associated with willingness to vaccinate24. Fears relating to the safety of the vaccine could also grow due to the relative speed of vaccine development or because vaccinating is now a reality rather than a hypothetical. Online misinformation could also play a role in shaping vaccination beliefs.

Despite these limitations, this study greatly extends existing research on both COVID-19 vaccine intentions and – more broadly – on the spatial resolution obtained for studies estimating nations’ vaccination beliefs or intentions^28,29^. By virtue of a more granular sub-national modelling approach, estimates are derived at regional scales consistent with those relevant for local policymaking or for improving epidemiological projections of COVID-19 mortality in the UK^30^. UK policymakers will need to be prepared to address vaccine concerns within the communities and regions identified in this study.

## Methods

### Data collection

Between 24 September and 14 October 2020, a cross-sectional online survey (see Supplementary Materials) probing acceptance of a novel COVID-19 vaccine was administered to 17,684 UK residents aged 18 and over. Informed consent was obtained from all respondents before the survey commenced. During data collection, quality control procedures resulted in the removal of 864 respondents (see *Methods*). The initial sample size was chosen to maximise the number of observations within each of the sub-national regions: this study has approximately 100 observations for each of the 174 sub-national regions, which far exceeds sample sizes used in similar research^31^. Respondent quotas were set according to national demographic distributions for sex, age, and sub-national region (the second level of the Nomenclature of Territorial Units for Statistics, or ‘NUTS2’, see https://www.ons.gov.uk/methodology/geography/ukgeographies/eurostat accessed 25 November 2020) and which were re-adjusted based on the removal of respondents through the ongoing quality control checks during data collection. These quotas ensured a geographic spread of respondents across the UK, between the sexes, and across all age groups. All respondents were recruited via an online panel by ORB (Gallup) International (www.orb-international.com) and informed consent was obtained before respondents participated.

The initial sample size was chosen to maximise the number of observations within each of the sub-national regions: this study has approximately 100 observations for each of the 174 sub-national regions, which far exceeds sample sizes used in similar research^31^. Respondent quotas were set according to national demographic distributions for sex, age, and sub-national region (the second level of the Nomenclature of Territorial Units for Statistics, or ‘NUTS2’, see https://www.ons.gov.uk/methodology/geography/ukgeographies/eurostat accessed 25 November 2020) and which were re-adjusted based on the removal of respondents through the ongoing quality control checks during data collection. These quotas ensured a geographic spread of respondents across the UK, between the sexes, and across all age groups. All respondents were recruited via an online panel by ORB (Gallup) International (www.orb-international.com) and informed consent was obtained before respondents participated.

The response variable is whether a respondent would accept a COVID-19 vaccine: “*If a new coronavirus (COVID-19) vaccine became available, would you accept the vaccine for yourself?*”, with responses on a four-point ordinal scale: “*yes, definitely*”, “*unsure, but leaning towards yes*”, “*unsure, but leaning towards no*”, or “*no, definitely not*”. The rationale behind this choice of responses is to elicit an explicit vaccination intent rather than provide a continuous or Likert scale, from which the intent to vaccinate may be less clear.

Covariate data are the socio-demographic traits collected for each individual and were chosen to align with the latest UK census: sex, age, highest educational attainment, religious affiliation, ethnicity, employment status, primary language, and outer postcode. Respondent’s outer postcode was used to map respondents to one of 174 third level NUTS regions (NUTS3). The maximum number of surveys conducted in a NUTS3 region is 293 (Hertfordshire) and the minimum is 16 (Mid and East Antrim). The mean number of responses per NUTS3 unit is 96.7 (with standard deviation 52.1) and the median is 85. A breakdown of the number of individuals surveyed by socio-demographic characteristic is found in Supplementary Materials, figure S1 and the survey counts for each NUTS3 region can be found in the supplementary data file.

### Multilevel regression and poststratification

Multilevel regression and poststratification (MRP) is used to estimate opinions aggregated at sub-national regions from survey data collected at the national level, via partial pooling of information between these national and sub-national scales^32^. This pooling of information between the two levels is a compromise between estimates derived via a total aggregation of data (to estimate national trends only) and estimates via complete disaggregation (that is, estimating regional trends only). The former suffers from a loss of information at the regional level while the latter suffers from possible low data counts and the loss of statistical power. More pooling of information will occur in regions with low relative numbers of surveyed individuals and less pooling in regions with high relative counts.

In brief (and relating specifically to this study), the first step of MRP is to conduct a multilevel regression to estimate, for each stratum (that is, a possible combination of individual characteristics) and for each region, the probability of COVID-19 vaccine acceptance. The second step is to reweight (post-stratify) these strata probabilities by the frequency with which a given strata appears in a population. In this study individual-level UK census data is used to perform the reweighting.

### Part 1: Multilevel regression

Individual intent to accept a COVID-19 vaccine is specified as *y*_*ij*_ ∈ {1,2,3,4}, where 1 = “no, definitely not”, 2 = “unsure, but leaning towards no”, 3 = “unsure, but leaning towards yes”, and 4 = “yes, definitely” and 1 < 2 < 3 < 4. Here, *j* =1, …, 174 is one of the *J* =174 third National Territorial Units for Statistics (NUTS3) regions in the UK, and *i* =1, …, *n*_*j*_, where *n*_*j*_ is the number of individuals surveyed in region *j*. ∑_*j*_ *n*_*j*_ =16,820 is the total number of respondents in the survey. A breakdown of the number of respondents in each region and a summary of their socio-econo-demographic status is given in the supplementary data file.

Intent to accept a COVID-19 vaccine is modelled as a multilevel ordinal regression with the proportional odds assumption^33^,

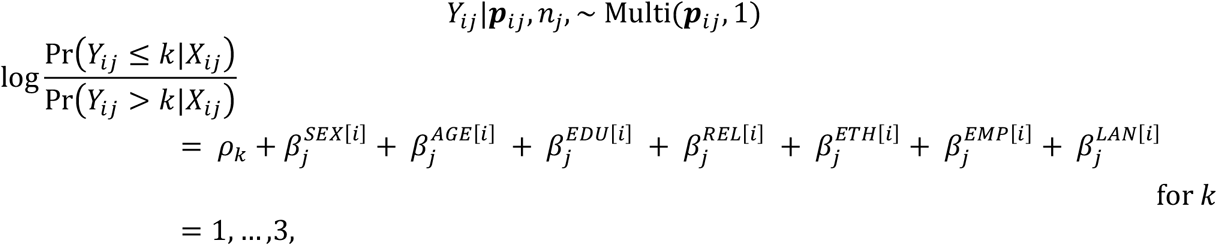

where 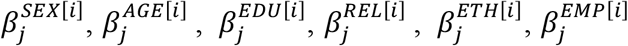, and 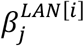 are the random-effect varying intercepts for sex, age, highest education level, religious affiliation, ethnicity, employment status, and primary language (respectively); *ρ*_*k*_ are probability threshold parameters; *k* ∈ {1,2,3,4} is the ordinal response category; ***p***_*ij*_ = [Pr(*Y*_*ij*_ =1), Pr(*Y*_*ij*_ = 2), Pr(*Y*_*ij*_ = 3), Pr(*Y*_*ij*_ = 4)]; and *X*_*ij*_ is the covariate data for individual *i* in region *j*. The baseline group for the regression corresponds to an individual who is male, aged 18-24, has an education level 1-3, is an atheist or agnostic, is White, works full-time, and speaks English or Welsh as their primary language.

In line with prior recommendations for variance components in hierarchical models ^32,34^, default weakly informative priors are chosen for the random-effects regression coefficients *β*^1^,

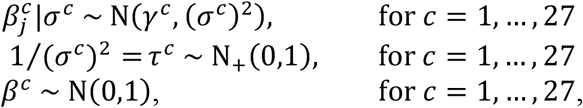

where *c* indexes the regression coefficients: excluding the threshold parameters, there are 27 fixed-effect parameters: 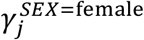, 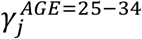, 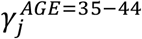, 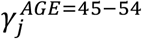, 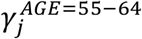, 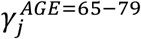, 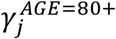, 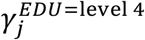, 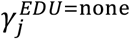, 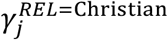, 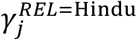, 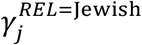, 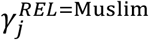, 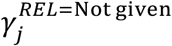, 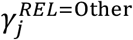, 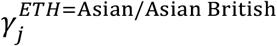, 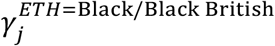, 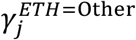, 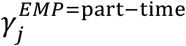, 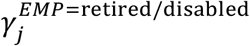, 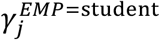, 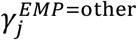, 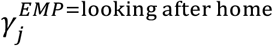, 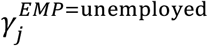, 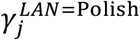, and 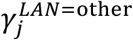. (And, thus, a total of 27 × 174 = 4,698 random effect parameters.)

### Part 2: Post-stratification

There are *S* = 30,870 socio-econo-demographic strata (two sexes × seven age groups × three education levels × seven affiliations for religion × five ethnicity groupings × seven employment statuses × three languages). Denoting the posterior probabilities of COVID-19 vaccination intent for each stratum *s* =1, …, *S* and NUTS3 region *j* =1, …, *J* as *θ*_*sjk*_ (where, as a reminder, *k* ∈ {1,2,3,4} denotes the response), then the MRP estimate for the intent to vaccinate within each of the UK’s 174 NUTS3 regions is,

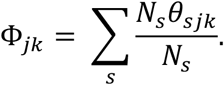

In the main text, this quantity Φ_*jk*_ is computed for *k* = 4 (“yes, [I] definitely [would accept a COVID-19 vaccine]”). Estimates are computed for those who are “unsure” (“unsure, but leaning towards yes” and “unsure, but leaning towards no” have been combined) and are shown in figure S1 in the supplementary materials.

### Model: Implementation and output

The multilevel regression model detailed above is implemented using JAGS version 4.3.0 (implemented via `rjags`^35^)and `R` version 4.0.3. 25,000 posterior samples (excluding the first 5,000 for model burn-in) was sufficient for successful convergence and all posterior draws were well-mixed. The posterior draws for the fixed effects are shown in figure S4 and all look visibly well-mixed and all except “other work status” (*p* = 0.04) have Geweke *p*-values above 0.05. There are too many posterior draws to plot for all random-effects, but we show posterior draws for the first UK NUTS3 region alphabetically (Hartlepool and Stockton-on-Tees) in figure S5 with a histogram of Geweke *p*-values for all model parameters (fixed effects, random effects, and variance components) to demonstrate universally good mixing and convergence in figure S3. In the computation of the Geweke statistic, the first 10% and final 50% of the posterior samples used for computation are used. Convergence of variance parameters is shown in figure S6. A slightly larger fraction of Geweke *p*-values fall below 0.05 than is expected by chance (0.082 compared to 0.05 by chance). Manual inspection of these chains revealed no cause for concern: chains showed no ill-mixing or convergence issues.

## Data Availability

Data for this study are not currently publicly available. The Author intends to make the data open access in early 2021.

## Funding

This project was funded by the Imperial College COVID-19 Response Fund.

## Competing interests

The Author is involved in Vaccine Confidence Project collaborative grants with GlaxoSmithKline outside the submitted work.

## Ethical Approval

Approval for this study was obtained via the Imperial College Research Ethics Committee on 24 July 2020 with reference 20IC6133 and European Union GDPR guidelines were followed throughout.

## Data and materials availability

All data used in this study will be made available at <GitHub URL to be inserted>.

## Footnotes

1 Instead of an noninformative N_+_(0,100) distribution over the standard deviation of hierarchical variance parameters ^34^, a weakly-informative N_+_(0,1) prior is placed over the *precision* of these parameters, which places 95% of *σ*^*c*^‘s prior mass between 0.54 and 4.05.

## Notes

### Author Declarations

Approval for this study was obtained via the Imperial College Research Ethics Committee on 24 July 2020 with reference 20IC6133.

### Summary of Updates

Model validation against observed first-dose uptake of COVID-19 vaccines in the UK has been added

