## supplementary materials for "Forecasting sub-national trends in COVID-19 vaccine uptake in the UK"

A. de Figueiredo, PhD[1,2†]

[1] Department of Infectious Disease Epidemiology, London School of Hygiene and Tropical Medicine, London, UK

[2] Department of Mathematics, Imperial College London, London, UK

[†]

These appendices contain a number of additional details and figures in support of Forecasted trends in COVID-19 vaccine acceptance across the UK: a large-scale cross-sectional spatial modelling study. The contents of these three appendices are detailed below.

### Appendix 1: Respondents' socio-econo-demographic background

The number (and percentage) of respondents within each socio-econo-demographic group (sex, age, highest education level, religious affiliation, ethnicity, employment status, and primary language) is shown in table S1. The number (and percentage) of each group replying to each of the four response options to "*If a new coronavirus (COVID-19) vaccine became available, would you accept it?*" is also shown.

| | | | | If a new coronavirus (COVID-19) vaccine became available, would you accept the vaccine for yourself? | | | | | | | |
|---|---|---|---|---|---|---|---|---|---|---|---|
| | Socio-econo-demographic | N | % | No, definitely not (N) | No, definitely not (%) | Unsure, but leaning towards no (N) | Unsure, but leaning towards no (%) | Unsure, but leaning towards yes (N) | Unsure, but leaning towards yes (%) | Yes, definitely (N) | Yes, definitely (%) |
| SEX | Female | 8682 | 51.6% | 788 | 9.1 | 1120 | 12.9 | 3071 | 35.4 | 3703 | 42.6 |
| | Male | 8138 | 48.4% | 560 | 6.9 | 699 | 8.6 | 2429 | 29.9 | 4450 | 54.7 |
| AGE | 18-24 | 1978 | 11.8% | 180 | 9.1 | 251 | 12.7 | 753 | 38.1 | 794 | 40.1 |
| | 25-34 | 2934 | 17.4% | 352 | 12 | 414 | 14.1 | 1022 | 34.8 | 1146 | 39.1 |
| | 35-44 | 3027 | 18.0% | 298 | 9.8 | 391 | 12.9 | 1042 | 34.4 | 1296 | 42.8 |
| | 45-54 | 3015 | 17.9% | 231 | 7.7 | 351 | 11.6 | 1025 | 34 | 1408 | 46.7 |
| | 55-64 | 2528 | 15.0% | 164 | 6.5 | 226 | 8.9 | 789 | 31.2 | 1349 | 53.4 |
| | 65-79 | 3150 | 18.7% | 117 | 3.7 | 176 | 5.6 | 826 | 26.2 | 2031 | 64.5 |
| | 80+ | 188 | 1.1% | 6 | 3.2 | 10 | 5.3 | 43 | 22.9 | 129 | 68.6 |
| highest EDUCATION | Level 1-3 | 7291 | 43.3% | 629 | 8.6 | 823 | 11.3 | 2476 | 34 | 3363 | 46.1 |
| | Level 4 | 7057 | 42.0% | 453 | 6.4 | 712 | 10.1 | 2221 | 31.5 | 3671 | 52 |
| | None/Other | 2472 | 14.7% | 266 | 10.8 | 284 | 11.5 | 803 | 32.5 | 1119 | 45.3 |
| RELIGIOUS affiliation | Atheist or agnostic | 5076 | 30.2% | 362 | 7.1 | 521 | 10.3 | 1692 | 33.3 | 2501 | 49.3 |
| | Christian | 8279 | 49.2% | 598 | 7.2 | 831 | 10 | 2549 | 30.8 | 4301 | 52 |
| | Hindu | 177 | 1.1% | 6 | 3.4 | 13 | 7.3 | 62 | 35 | 96 | 54.2 |
| | Jewish | 143 | 0.9% | 7 | 4.9 | 11 | 7.7 | 38 | 26.6 | 87 | 60.8 |
| | Muslim | 534 | 3.2% | 66 | 12.4 | 80 | 15 | 209 | 39.1 | 179 | 33.5 |
| | Not given | 1159 | 6.9% | 134 | 11.6 | 164 | 14.2 | 439 | 37.9 | 422 | 36.4 |
| | other religion | 1452 | 8.6% | 175 | 12.1 | 199 | 13.7 | 511 | 35.2 | 567 | 39 |
| ETHNICITY | Asian/Asian British | 845 | 5.0% | 65 | 7.7 | 103 | 12.2 | 336 | 39.8 | 341 | 40.4 |
| | Black/Black British | 413 | 2.5% | 81 | 19.6 | 88 | 21.3 | 113 | 27.4 | 131 | 31.7 |
| | Mixed | 321 | 1.9% | 41 | 12.8 | 40 | 12.5 | 107 | 33.3 | 133 | 41.4 |
| | other ethnicity | 315 | 1.9% | 52 | 16.5 | 51 | 16.2 | 112 | 35.6 | 100 | 31.8 |
| | White | 14926 | 88.7% | 1109 | 7.4 | 1537 | 10.3 | 4832 | 32.4 | 7448 | 49.9 |
| EMPLOYMENT status | full-time | 7326 | 43.6% | 641 | 8.8 | 801 | 10.9 | 2371 | 32.4 | 3513 | 48 |
| | looking after home / family | 925 | 5.5% | 89 | 9.6 | 133 | 14.4 | 345 | 37.3 | 358 | 38.7 |
| | other | 105 | 0.6% | 23 | 21.9 | 12 | 11.4 | 37 | 35.2 | 33 | 31.4 |
| | part-time | 2942 | 17.5% | 238 | 8.1 | 394 | 13.4 | 1079 | 36.7 | 1231 | 41.8 |
| | retired / disabled | 3834 | 22.8% | 192 | 5 | 285 | 7.4 | 1033 | 26.9 | 2324 | 60.6 |
| | student | 844 | 5.0% | 54 | 6.4 | 92 | 10.9 | 340 | 40.3 | 358 | 42.4 |
| | unemployed | 844 | 5.0% | 111 | 13.2 | 102 | 12.1 | 295 | 35 | 336 | 39.8 |
| primary LANGUAGE | English or Welsh | 15656 | 93.1% | 1179 | 7.5 | 1637 | 10.5 | 5090 | 32.5 | 7750 | 49.5 |
| | Other | 1026 | 6.1% | 138 | 13.4 | 163 | 15.9 | 360 | 35.1 | 365 | 35.6 |
| | Polish | 138 | 0.8% | 31 | 22.5 | 19 | 13.8 | 50 | 36.2 | 38 | 27.5 |

***Table S1.*** **Socio-econo-demographic breakdown of the 16,820 survey participants and their intent to accept a COVID-19 vaccine**

### Appendix 2: Supplementary figures

##### Mapping COVID-19 vaccine acceptance: unsure about accepting a COVID-19 vaccine

The estimated proportion of respondents in each of the UK's 174 NUTS3 regions who are unsure about whether they would accept a COVID-19 vaccine ('unsure, but leaning towards yes" or "unsure, but leaning towards no") is shown in figure S1 below. Estimates are derived via multilevel regression and poststratification as described in the main text and Appendix 3.

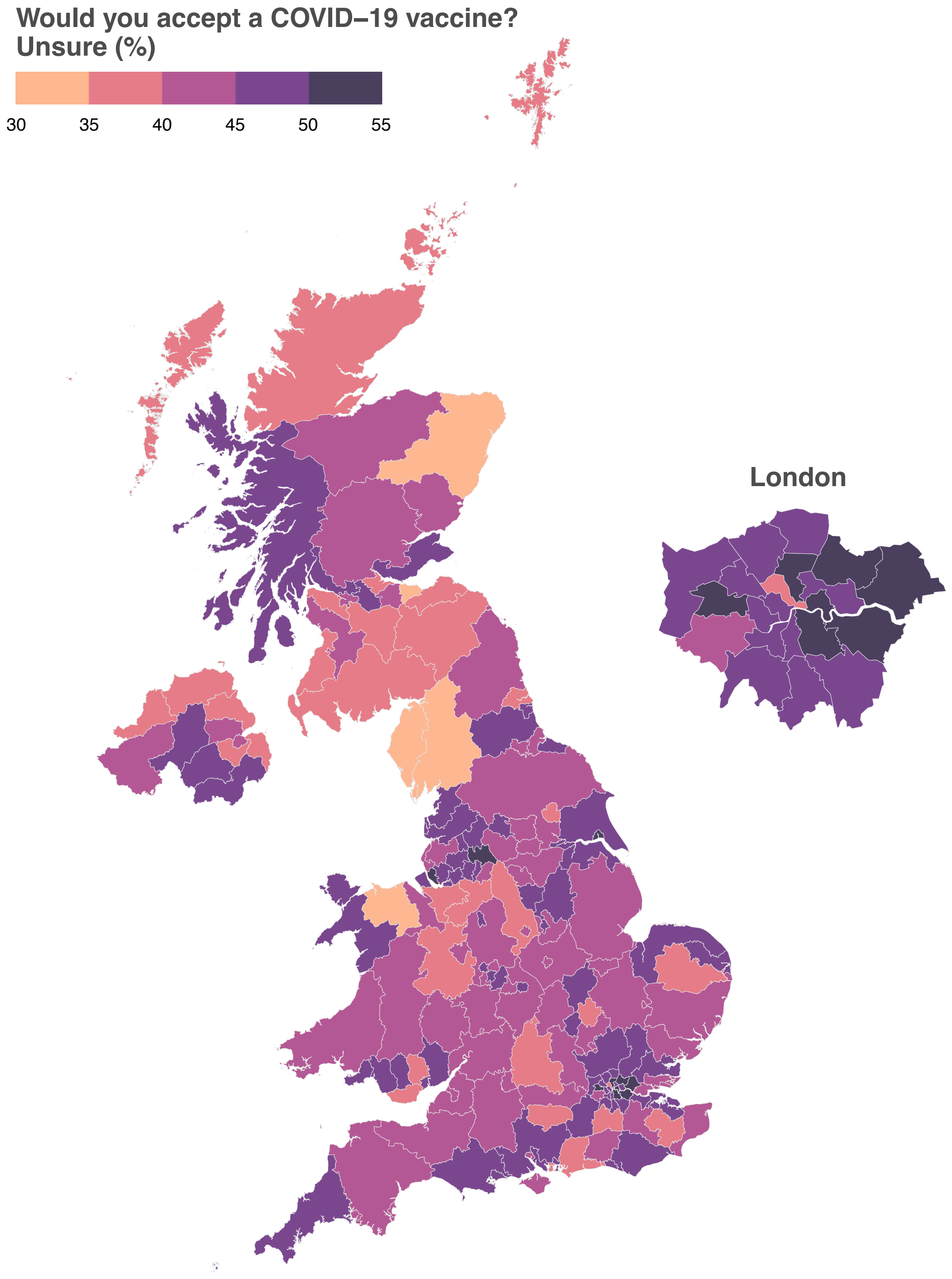

***Figure S1*. Regional level MRP estimates of the proportion of each UK region who are unsure about accepting a COVID-19 vaccine**

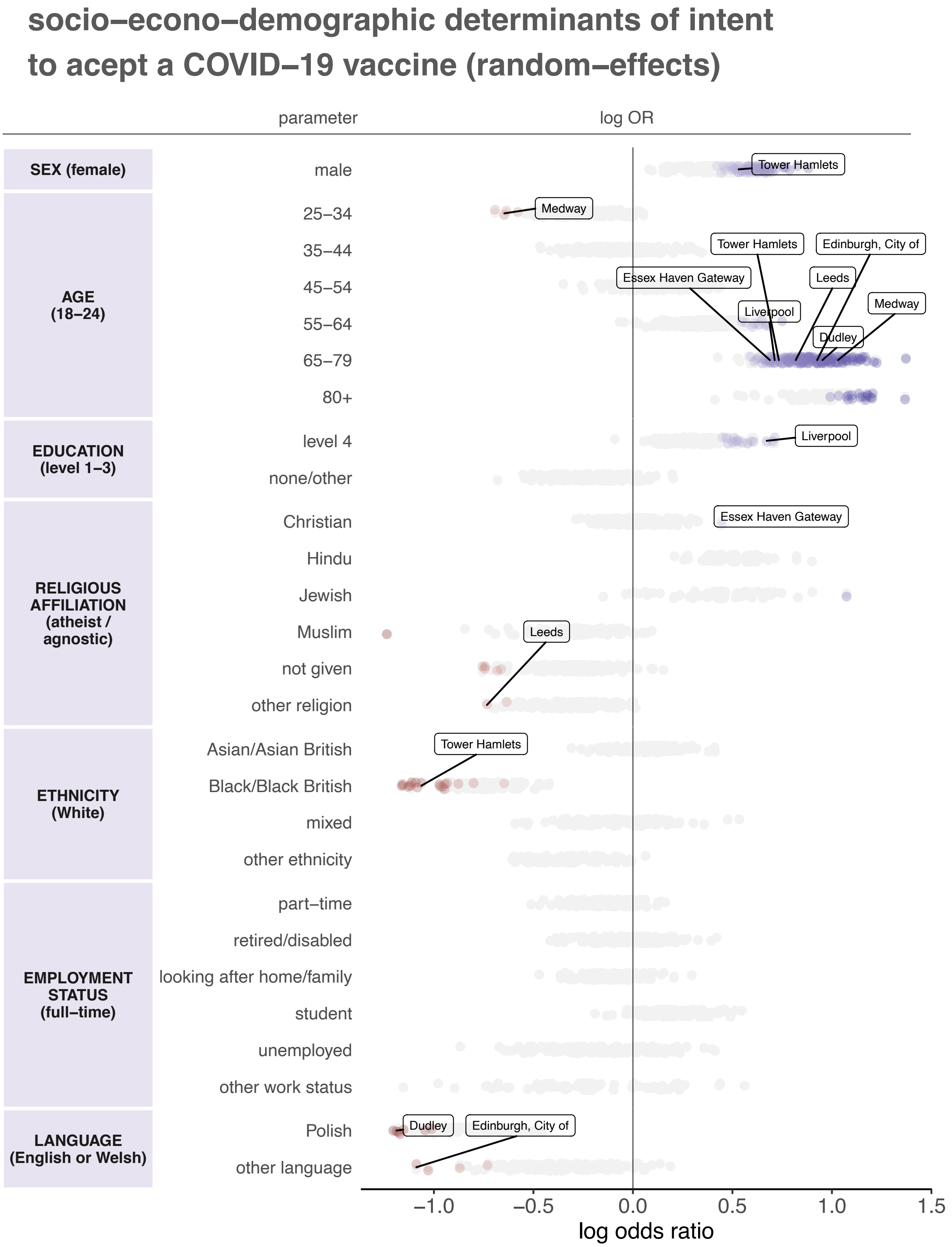

*Figure S2.* **Multilevel regression random effects.** All random effect parameters from the multilevel regression are shown in grey. Parameter log odds ratios are plotted with corresponding 95% HPDIs and coloured by effect size, where blues (reds) denote a positive (negative) association between the factor and intent to accept a vaccine relative to the baseline group (which is provided in parentheses on the left column). The darker the colour the stronger the association. All parameters whose 95% HPDI excludes zero are coloured. A selection of sub-national regions are shown, see supplementary data file (random effects) for all inferred random-effect parameters with HPDIs for all 174 regions.

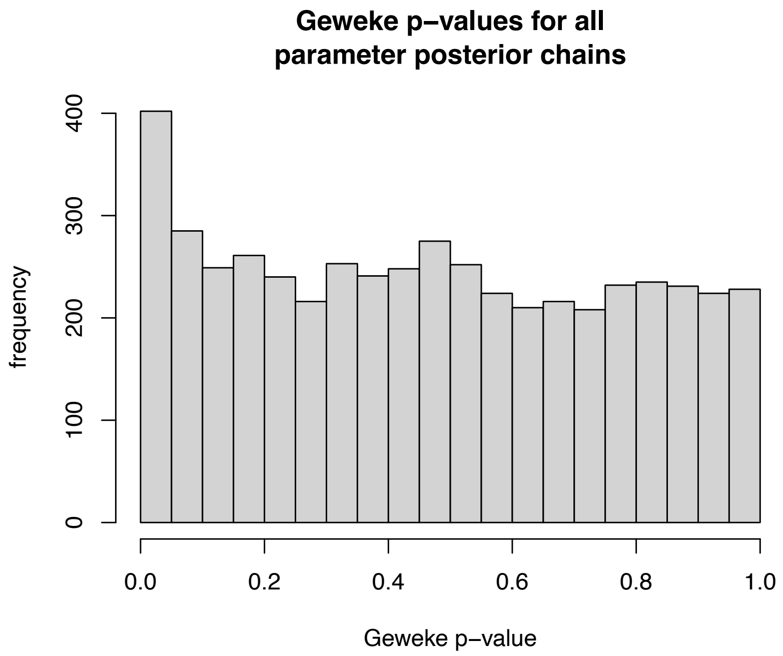

***Figure S3*. Distribution of Geweke *p*-values for all parameter posterior chains.**

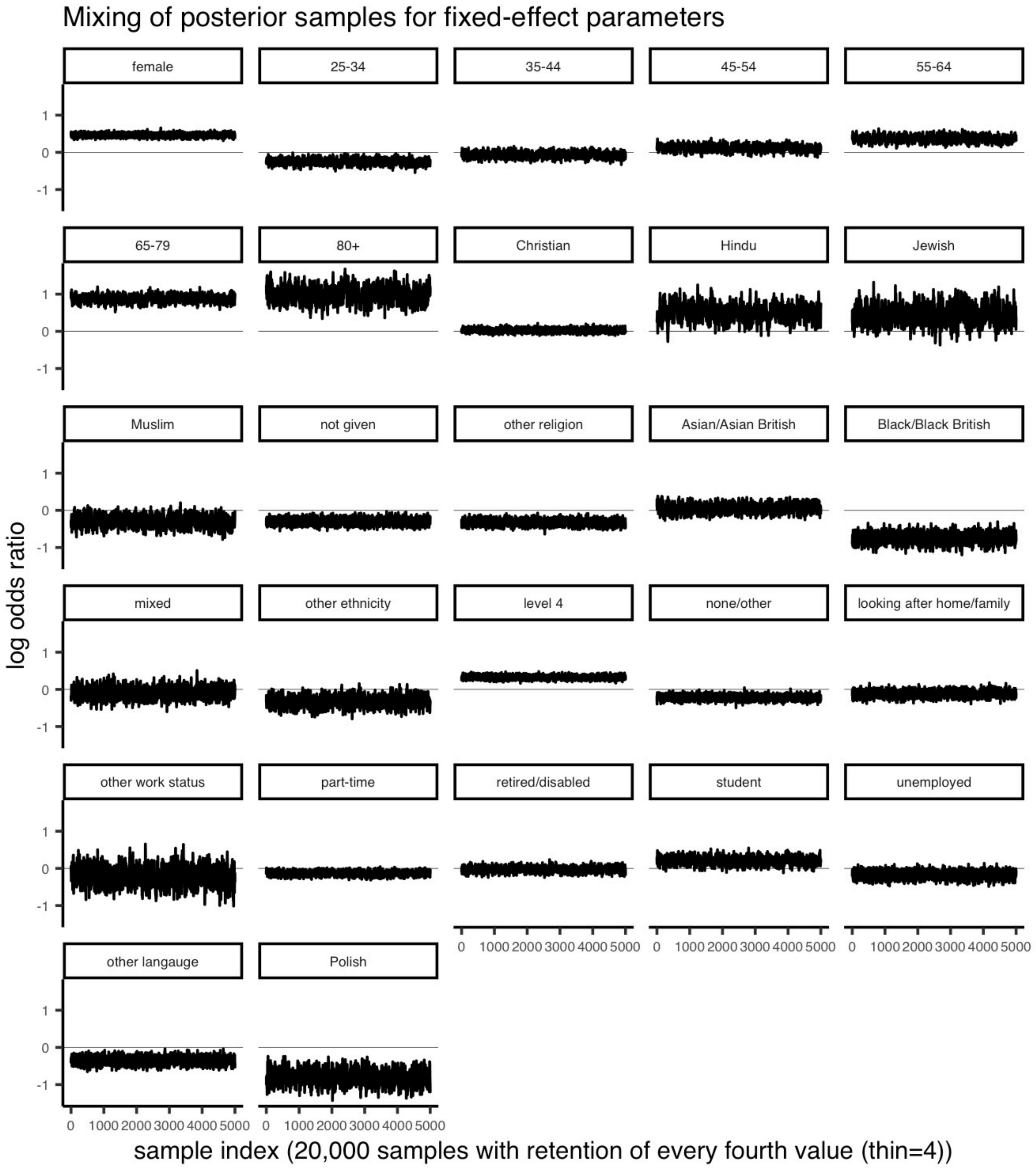

***Figure S4*** **Mixing and convergence of the fixed-effect parameters in the multilevel regression model**

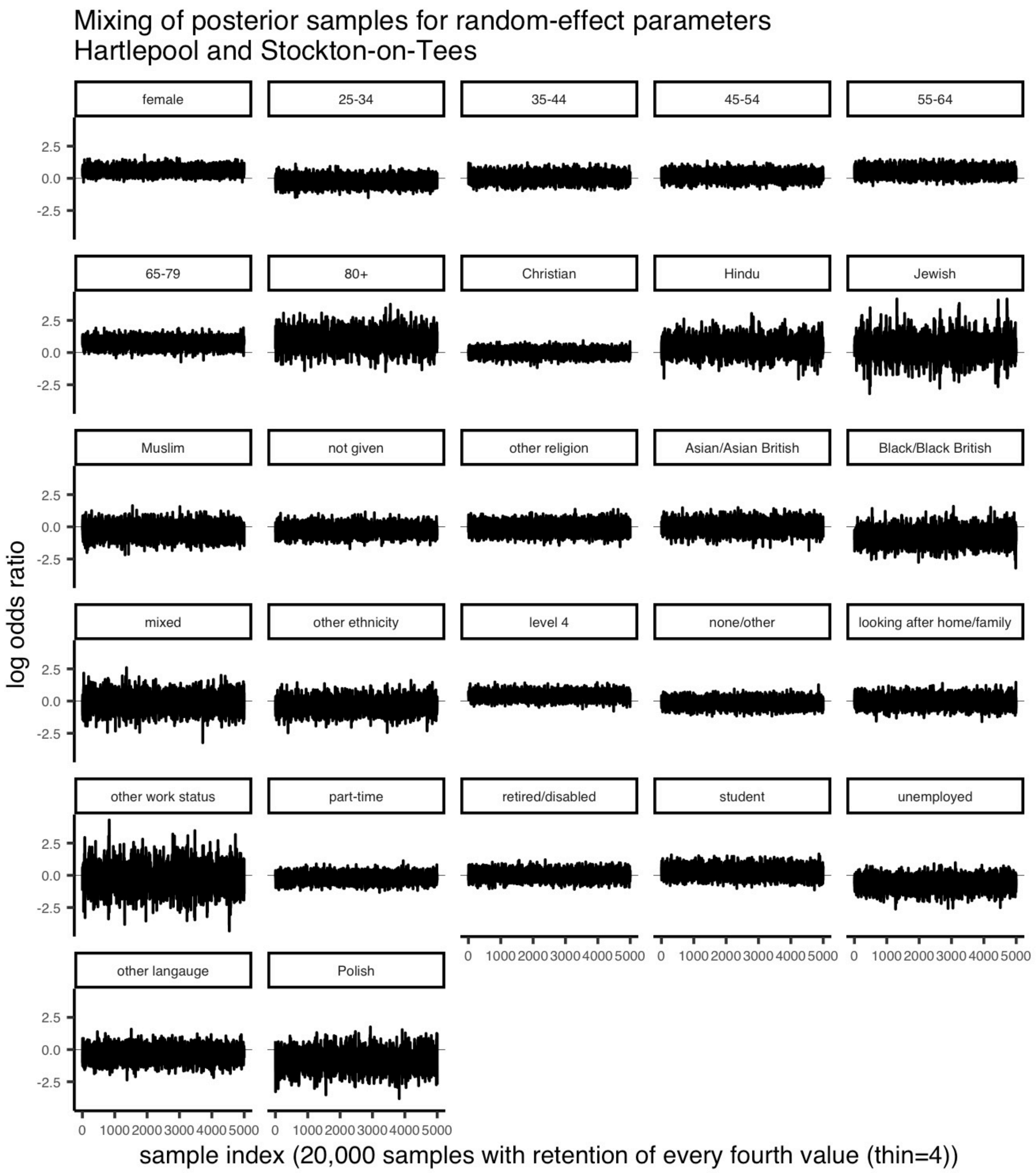

***Figure S5*** **Mixing and convergence of the random-effect parameters for Hartlepool and Stockton-on-Tees in the multilevel regression model.**

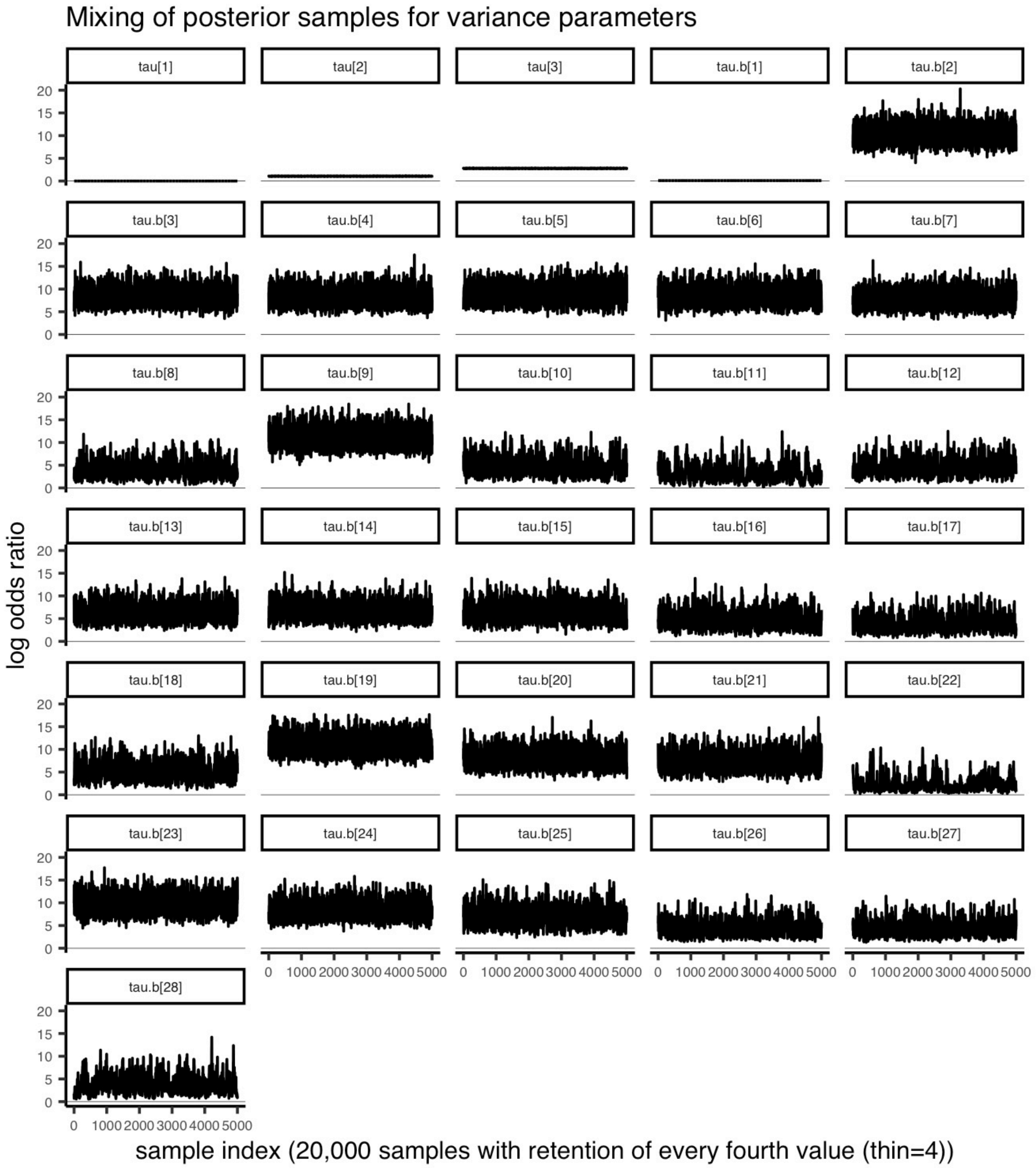

***Figure S6* Mixing and convergence of the variance (tau.b[1:28]) and threshold (tau[1:4]) parameters for in the multilevel regression model. Note that for model identifiability, tau[1] = 0 is fixed.**
